# A higher ratio of green spaces means a lower racial disparity in severe acute respiratory syndrome coronavirus 2 infection rates: A nationwide study of the United States

**DOI:** 10.1101/2020.11.11.20228130

**Authors:** Yi Lu, Long Chen, Xueming Liu, Yuwen Yang, Wenyan Xu, Chris Webster, William C. Sullivan, Bin Jiang

**Affiliations:** Department of Architecture and Civil Engineering, College of Engineering, City University of Hong Kong, Hong Kong SAR; Virtual Reality Lab of Urban Environments and Human Health, HKUrbanLabs, The University of Hong Kong, Hong Kong SAR; Division of Landscape Architecture, Department of Architecture, The University of Hong Kong, Hong Kong SAR; HKUrbanLabs, Faculty of Architecture, The University of Hong Kong, Hong Kong SAR; Smart, Healthy Communities initiative, University of Illinois at Urbana-Champaign, U.S.A.

**Author notes:** Corresponding author: Postal Address: 614 Knowles Building, The University of Hong Kong, Pokfulam Road, Hong Kong SAR.

**Keywords:** racial disparity, health disparity, SARS-CoV-2, COVID-19, green space, pathway

## Abstract

There is striking racial disparity in the severe acute respiratory syndrome coronavirus 2 (SARS-CoV-2) infection rates in the United States. We hypothesize that the disparity is significantly smaller in areas with a higher ratio of green spaces at the county level. This study used the 135 most urbanized counties across the United States as sample sites. County level data on the SARS-CoV-2 infection rates of black and white individuals in each county were collected. The ratio of green spaces by land-cover type at the county level was calculated from satellite imagery. An ecological hierarchical regression analysis measured cross-sectional associations between racial disparity in infection rates and green spaces, after controlling for socioeconomic, demographic, pre-existing chronic disease, and built-up area factors. We found significantly higher infection rate among black individuals compared to white individuals. More importantly, a higher ratio of green spaces at the county level is significantly associated with a lower racial disparity in the SARS-CoV-2 infection rate. Further, we identified four green space factors that have significant negative associations with the racial disparity in SARS-CoV-2 infection rates, including open space in developed areas, forest, shrub and scrub, and grassland and herbaceous. We suggest that green spaces are an equalizing salutogenic factor, modifying infection exposure.

**Highlights:**

- The first study to identify significant relationships between green spaces and the racial disparity of SARS-CoV-2 infection rates.
- A nationwide study of the 135 most urbanized counties of the United States.
- A within-subject study: The black-white racial disparity of SARS-CoV-2 infection rates was measured within each county.
- A higher ratio of green spaces in a county is associated with a lower racial disparity of SARS-CoV-2 infection rates after controlling for socio-economic, demographic, pre-existing chronic disease, and built-up area factors.
- Four green space factors are significantly associated with a lower racial disparity of SARS-CoV-2 infection rates.

## Introduction

Racial disparity in health is a significant problem in many countries and can lead to social conflicts, economic crises, and loss of life^1, 2^. The black–white health disparity in the United States is a representative example in developed economies and is unsurprisingly manifest in the ongoing coronavirus disease 2019 (COVID-19) pandemic^3, 4, 5, 6^. COVID-19 results from infection with severe acute respiratory syndrome coronavirus 2 (SARS-CoV-2), and the SARS-CoV-2 infection rate in black individuals is significantly higher, proportionally, than that in white individuals^4, 7, 8^. Although studies of the COVID-19 pandemic are quickly accumulating, only a small portion have examined racial disparity in infection rates^9, 10, 11, 12^. Most of these studies focused on relationships between socioeconomic, demographic, or pre-existing chronic disease factors and racial disparity in SARS-CoV-2 infection rates ^4, 13, 14^. None of the studies have directly examined effects of urban environmental factors. Evidence suggests that green spaces may have positive and independent effects on reducing the racial disparity in various long-term health outcomes ^15, 16, 17, 18, 19, 20, 21^. Yet, to our knowledge, there is no evidence to support green space’s effect on reducing racial disparity in SARS-CoV-2 infection rates. This lack of knowledge may mean missing opportunities to slow down the COVID-19 pandemic, and to moderate future epidemics by through urban greening. Our nationwide study is an initial ecological cross-sectional study aiming to establish a county-level relationship between black–white racial disparity in SARS-CoV-2 infection rates and the amount and type of green spaces in the United States.

### A new trend: Significant environmental effects on racial disparity in health

Many studies have explored potential mechanisms that maintain and exacerbate general racial disparity in health outcomes to identify effective policies to address such inequities. Much of the racial disparity seems to be influenced by differences in socioeconomic, demographic, and pre-existing chronic disease factors^3, 4, 14^. Still, disparities remain significant after controlling for these factors. For example, the life expectancies of higher socioeconomic status (SES) black and white individuals are 7.1 and 6.8 years longer, respectively, than those of their lower SES counterparts^22, 23^. Life expectancy of black people is at least three years less than that of white people with a comparable income level, at every level of income^23^. Racial disparity is also pronounced in birth outcomes; for example, the infant mortality rate for college-educated black women is 2.5 times higher than that for college-educated white women^23, 24^. These studies suggest that persistent racial health disparities are due, in part, to other types of systemic disparities between races.

After controlling for SES factors, an under-studied reason for racial disparity in health outcomes is built environment disparity ^25^. People of different races, especially black and white people in the USA, have lived in neighborhoods with distinct environmental qualities^26^. Neighborhoods that are the long-term home of more black people have, on average, fewer green spaces for physical activity, lower accessibility to healthy food, and lower traffic or crime-related safety^27, 28, 29, 30^. These findings have prompted studies in new directions. Researchers across disciplines have shifted from investigating exclusively social, economic, and educational interventions, and are now also investigating physical environmental interventions as a means to alleviate persistent racial-disparities^19, 29, 31^.

### Positive effects of green spaces on human health

The significantly positive effects that green spaces have on human health have drawn considerable attention from researchers, public health professionals, and governmental officers over the past decades. A growing number of studies have measured these effects at the regional^19, 32^, municipal^33^, neighborhood^34^, and property-site^35^ levels. Consensus has emerged that green spaces can influence human health through five major pathways^36^, namely attention restoration^37^, stress reduction^38, 39^, enhanced social cohesion and social capital^40^, increased physical activity^41^, and supply of ecological products and services^42, 43, 44^. These pathways have been demonstrated across different geographic, social, and cultural contexts^45, 46, 47^.

### Green spaces reduce racial disparity in health: Circumstantial evidence and knowledge gaps

One line of research might be highly relevant to racial disparity in SARS-CoV-2 infection rates: green spaces can mitigate health disparities among populations with distinctively different SESs. A series of observational and survey studies found that a greater amount of green space in public housing communities was significantly correlated with a higher level of self-discipline^48^, a lower level of mental fatigue^49, 50^, a higher level of social activity and cohesion^51^, and a lower level of stress and aggressive behavior^52^ in residents. A nationwide study in the UK revealed that the association between income level and all-cause mortality and circulatory disease mortality at the neighborhood level can be moderated by the amount of green space in a neighborhood^19^. The study suggested that more exposure to green space can reduce the health disparity caused by income disparity. The supply of public green spaces can reduce health disadvantages due to obesity and obesity-related illnesses in residents of low SES, by encouraging physical exercise^21^. Some citywide experimental studies found that the supply of green open space in low-income neighborhoods can significantly reduce crime, violence, stress, and fear, and increase daily exercise in those neighborhoods^53, 54^.

Many of these studies have examined the relationships between the supply of green spaces and health disparities in long-term health between populations with distinctively different SESs^55, 56^. While SES disparity is relevant to racial disparity, it is not fully representative of it^57^. Furthermore, the potential for exposure to green spaces to attenuate racial disparity in the risk of infection by airborne viruses, such as SARS-CoV-2, has not been investigated. This is a critical knowledge gap that may limit our opportunities to moderate the current pandemic and epidemics in the future.

### Hypothesis

We hypothesized that the black–white racial disparity in the USA’s SARS-CoV-2 infection rates is significantly less in urbanized counties with a higher ratio of various types of green spaces.

## Methods

### Study design

We compared the black–white disparity in SARS-CoV-2 infection rates in populations living in urbanized counties of the United States that have different amounts of green spaces, while adjusting for potential confounding factors of these counties. The infection data were retrieved on July 10, 2020. We adopted a within-subject (within-county) research design with a representative sample across the country. Black–white disparity was measured as the difference in the infection rate of black and white individuals in the same county. This allowed us to largely remove the bias caused by between-subject (between-county) factors that may lead to the uneven spread of SARS-CoV-2 in different counties, such as the distance to COVID-19 epicenters, climate, or local government policies. The study is cross-sectional in the sense that it does not analyze across time and ecological in the sense that it uses data aggregated at spatial unit of analysis (county).

### Study areas

The United States is among the countries most severely impacted by COVID-19. It also has severe black–white racial inequalities in SARS-CoV-2 infection rates. Our study used counties as the basic unit of analysis, which are the fundamental administrative unit in the United States. There are a total of 3,142 counties or county-equivalent areas in the United States.

The most urbanized counties were chosen because we assumed that the racial inequality in the SARS-CoV-2 infection rates is more pronounced in denser urban environments. First, we identified a list of 314 large cities with a population ≥ 100,000 in 2019. Second, we identified a total of 229 counties containing or overlapping these large cities. Third, counties without infection data for black and white people were excluded. A total of 135 counties were chosen as our study areas. The total population in these counties was 132,350,027, which comprised 40.3% of the total population in the United States. The large population size in our study areas ensures the generalizability of potential findings. We chose counties rather than cities as the unit of analysis because the infection data for black and white individuals were often unavailable at the city level.

### SARS-CoV-2 infection rates

The infection data were collected from the county health department portals on July 10, 2020. Only SARS-CoV-2 infections in black and white individuals were collected in this study. We calculated the SARS-CoV-2 infection rates for black and white people (numbers of cases per 100k) based on the total white and black populations in each county, respectively, which were retrieved from the 2019 census data^58^. The racial disparity in the SARS-CoV-2 infection rates was calculated as the difference between the infection rate in black individuals and the infection rate in white individuals in the same county.

### Green spaces

We used three datasets to assess green spaces, namely the National Land Cover Datasets in 2016 (NLCD 2016), the Tree Canopy Cover Datasets of the United States Forest Service (USFS), and NDVI.

The NLCD 2016 provides spatially explicit and reliable information on the national land-cover classification based on Landsat imagery at 30-meter resolution^59^. The overall agreement ranges from 71% to 97% between different land-cover classifications and reference data^59^. The NLCD 2016 has 16 types of land covers. We considered the following land-cover types with dominant natural elements: developed open space, deciduous forest, evergreen forest, mixed forest, shrub and scrub, grassland and herbaceous, pasture and hay, cultivated crops, woody wetlands, and emergent herbaceous wetlands (Appendix Table 2). Deciduous/evergreen/mixed forest were combined into a new land-cover denoted “forest”. The ratio of these green spaces over the total county area in each county was calculated.

The Tree Canopy Cover Dataset was derived from the NLCD by the USFS. It provides estimates of the percent tree canopy cover at 30-meter resolution. We aggregated the total tree-canopy cover estimates for each county and then divided it by the county area.

The NDVI is a widely used index that measures the quantity of green vegetation cover at the pixel level via remote sensing. We obtained the NDVI at a 30-meter resolution from the Google Earth Engine, which integrates Landsat 8 imagery in its cloud platform^60^. Four pixel-level average values were calculated for each county in each month from March to July in 2020, and we finally obtained the mean of four monthly NDVI values as a proxy for overall green spaces coverage during the research period.

However, we found that the land-cover dataset had high multicollinearity with two other measurements of green spaces. For instance, the NDVI and forest land-cover are strongly correlated (Pearson correlation coefficient: *r* = 0.64, *p* < 0.001), as are tree canopy and forest (*r* = 0.85, *p* < 0.001). Therefore, we only used measures from the land-cover dataset.

### Confounding factors

Previous studies have shown that demographic and socioeconomic characteristics are important predictors of SARS-CoV-2 infection risk or racial disparity in this risk^1, 2, 61^. From the 2019 census data, we obtained the population density, the female population ratio, the difference in black–white population, the difference in black–white older adults, household size, housing value, the rate of high school graduates or higher, the rate of households with broadband, median household income, the poverty rate, healthcare receipts data, travel time to work, the employment rate, and the number of firms for our study areas (Appendix Table 1)^58, 62^.

Pre-existing chronic disease is another indispensable dimension that might be connected with the racial disparity in SARS-CoV-2 infection rates^61^. Thus, county-level pre-existing chronic disease factors were collected from the 2016–2018 interactive heart disease and stroke data collated by the Centers for Disease Control and Prevention^63^. The pre-existing chronic disease factors that we used were the coronary heart disease death rate, the heart failure death rate, the diagnosed diabetes rate, and the obesity rate for each county (Appendix Table 1).

In addition, land-cover types of dominant built-up area were considered because they represent the level of urbanity of each county. Three land-cover types of built-up area from NLCD 2016 were included: developed low intensity, developed medium intensity, and developed high intensity (See detailed definitions in the Appendix Table 2). The ratio of these built-up areas over the total county area in each county was calculated as the same way that we calculated the ratio of green spaces in each county.

All confounding factors were represented by proportional variables (e.g., per 100 k, density, ratio, and median value) or the difference between black and white individuals at the county level. Therefore, they were consistent with the dependent variable.

### Statistical analysis

Three statistical analysis steps were performed. First, a paired *t*-test was used to examine whether there was a significant difference between SARS-CoV-2 infection rates in black and white people in the same county. Second, a variance inflation factor (VIF) test was used to remove potential multicollinearity among the independent variables. We adopted the threshold value of VIF = 4^64^. All of the factors with a VIF ≥ 4 were excluded from our models (Appendix Tables 1 & 2).

Third, hierarchical linear-regression models were used to examine the associations between the black–white difference in the SARS-CoV-2 infection rates and green space factors, while controlling for other confounding factors. The first model (Model 1) included the socioeconomic and demographic factors only. The second model (Model 2) additionally included pre-existing chronic disease factors. The last model (Model 3) additionally included green space factors, on the basis of Model 2.

All of the analyses were performed using R v4.0.2^65^. The model *R*^2^ values, adjusted *R*^2^ values, standardized coefficient (*β*) values, 95% confidence intervals, and *p*-values were reported.

### Results

Results are presented in three sections. First, we present the incidence of SARS-CoV-2 infections in the sample of 135 most urbanized counties in the United States and examine the extent to which there are differences in infection rates among black and white populations. Second, we report demographic and green space characteristics of the sampled counties. Third, we employ hierarchical linear modeling to examine the extent to which, after controlling for socioeconomic, demographic, pre-existing chronic disease, and built-up area factors, the ratio of green spaces are associated with black-white disparities in SARS-CoV-2 infection rates. Finally, we report four green space factors that have significant negative associations with the racial disparity in SARS-CoV-2 infection rates.

### Does a racial disparity in infection rates exist?

As of July 10, 2020, there were a total of 1,425,461 cases of SARS-CoV-2 infection in the 135 most urbanized counties in the United States, which accounted for 47% of the total cases of infection (3,038,325) in the United States (Figure 1). The 135 most urbanized counties were selected because they contain or overlap with 314 large cities with a population ≥ 100,000 in 2019 (see details in Methods). The county-level average infection rate for white individuals was 497 persons per 100,000 population, whereas the infection rate for black individuals was approximately twice this (988 persons per 100,000 population) (Figure 1 & Table 1). The average black–white difference in the infection rate was 447 persons per 100,000 population. White individuals had a higher infection rate than did black individuals in only 11 out of the 135 counties (Figure 2). As expected, a paired *t*-test revealed that the difference infection rates between black and white people is significant, with *t* (134) = 12.757 and *p* < 0.001 (Figure 3).

**Table 1.** Descriptive statistics for SARS-CoV-2 infection rates, socioeconomic and demographic factors, pre-existing chronic disease factors, and green space factors in the USA’s 135 most urbanized counties.

| Variable Categories | Variables | Min | Max | Mean | SD | Unit or Formula |
| --- | --- | --- | --- | --- | --- | --- |
| **Infection outcomes** | Black infection rate | 91.6 | 2,514.8 | 987.9 | 583.7 | Cases per 100k |
|  | White infection rate | 29.1 | 2,486.2 | 496.8 | 336.2 | Cases per 100k |
|  | Difference in black–white infection rates | -367.9 | 1,874.2 | 491.1 | 447.3 | Cases per 100k |
| **Socioeconomic and demographic factors** | Population density | 27.8 | 19,625.8 | 1,003.2 | 2,356.6 | Persons per square kilometer |
|  | Female population ratio | 0.479 | 0.533 | 0.511 | 0.010 | Female/ total population |
|  | Difference in black–white population | -0.878 | 0.481 | -0.407 | 0.274 | Black-to-white population ratio |
|  | Difference in black–white older adults | -0.201 | 0.050 | -0.078 | 0.041 | Black older adults-to-white older adults population ratio |
|  | Household size | 2.1 | 3.3 | 2.6 | 0.3 | Persons per household |
|  | Households with broadband | 0.666 | 0.917 | 0.818 | 0.056 | Ratio |
|  | Median household income | 38,085 | 117,374 | 62,484 | 16,126 | USD |
|  | Healthcare receipts | 222,956 | 2,129,326 | 771,836 | 310,816 | 1000 USD per 100k |
|  | Number of firms | 5,000 | 19,365 | 8,595 | 2,076 | Number of firms per 100k |
| **Pre-existing chronic disease factors** | Coronary heart disease death rate | 45.3 | 144.9 | 85.1 | 22.0 | Cases per 100k |
|  | Heart failure death rate | 34.4 | 170.3 | 89.1 | 24.6 | Cases per 100k |
|  | Diagnosed diabetes rate | 0.046 | 0.140 | 0.090 | 0.018 | Cases/ total population |
| **Green space factors** | Developed open space | 0.005 | 0.289 | 0.102 | 0.064 | Developed open space area/ county area |
|  | Forest | 0.001 | 0.738 | 0.172 | 0.163 | Forest area/ county area |
|  | Shrub and scrub | 0.000 | 0.829 | 0.082 | 0.177 | Shrub and scrub area/ county area |
|  | Grassland and herbaceous | 0.001 | 0.498 | 0.061 | 0.093 | Grassland and herbaceous area/ county area |
|  | Pasture and hay | 0.000 | 0.485 | 0.069 | 0.092 | Pasture and hay area/ county area |
|  | Cultivated crops | 0.000 | 0.719 | 0.124 | 0.168 | Cultivated crops area/ county area |
|  | Woody wetlands | 0.000 | 0.453 | 0.050 | 0.078 | Woody wetlands area/ county area |
|  | Emergent herbaceous wetlands | 0.000 | 0.605 | 0.026 | 0.080 | Emergent herbaceous wetlands area/ county area |

Note: Min = minimum; Max = maximum; SD = standard deviation; Per 100k = 100,000 population; USD = United States Dollar.

**Figure 1.**
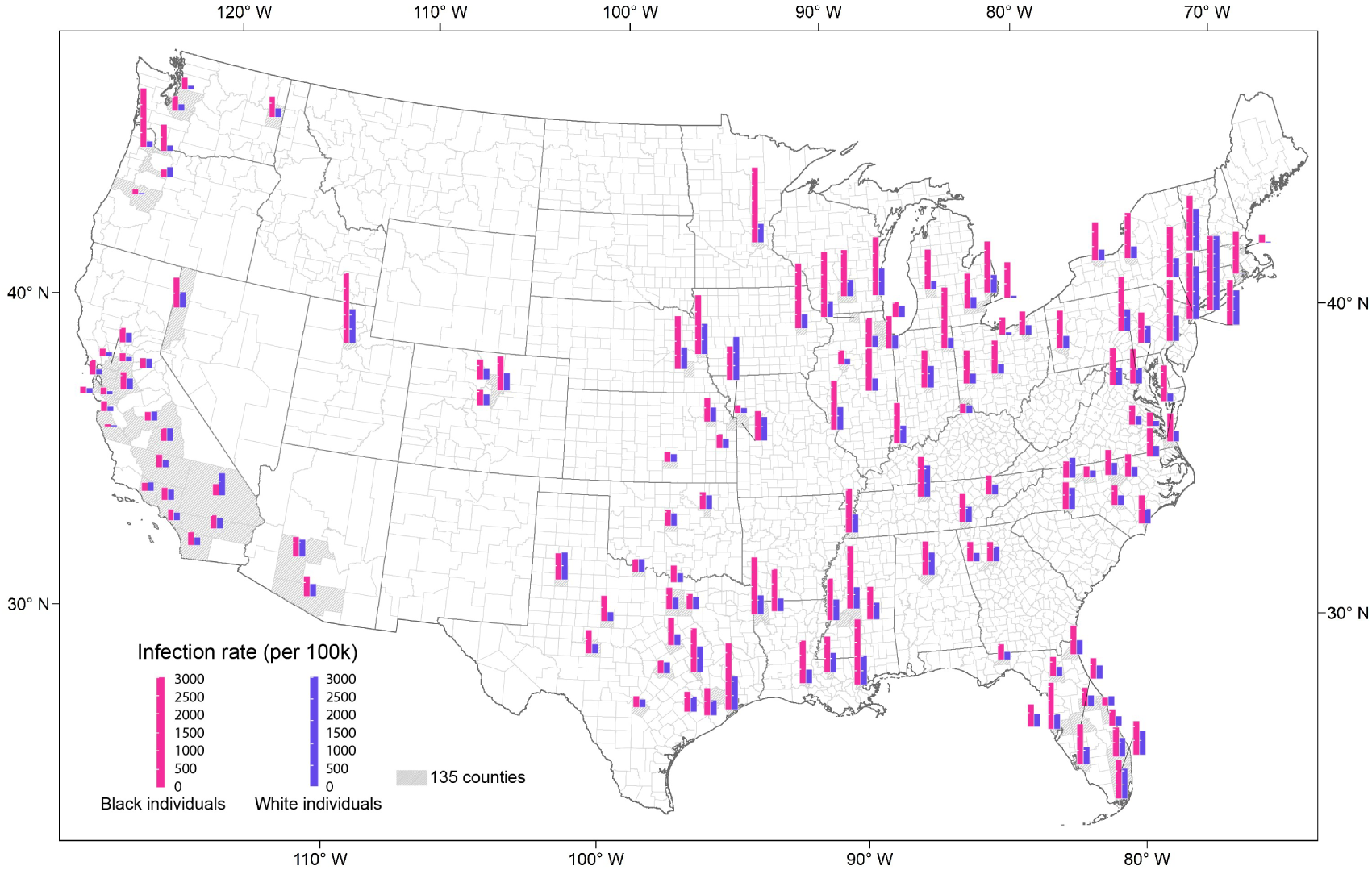
SARS-CoV-2 infection rates of black and white individuals in the 135 most urbanized counties of the United States. Pink column indicates infection rate for black people; blue column indicates infection rate for white people. The height of columns indicates the magnitude of infection rate.

**Figure 2.**
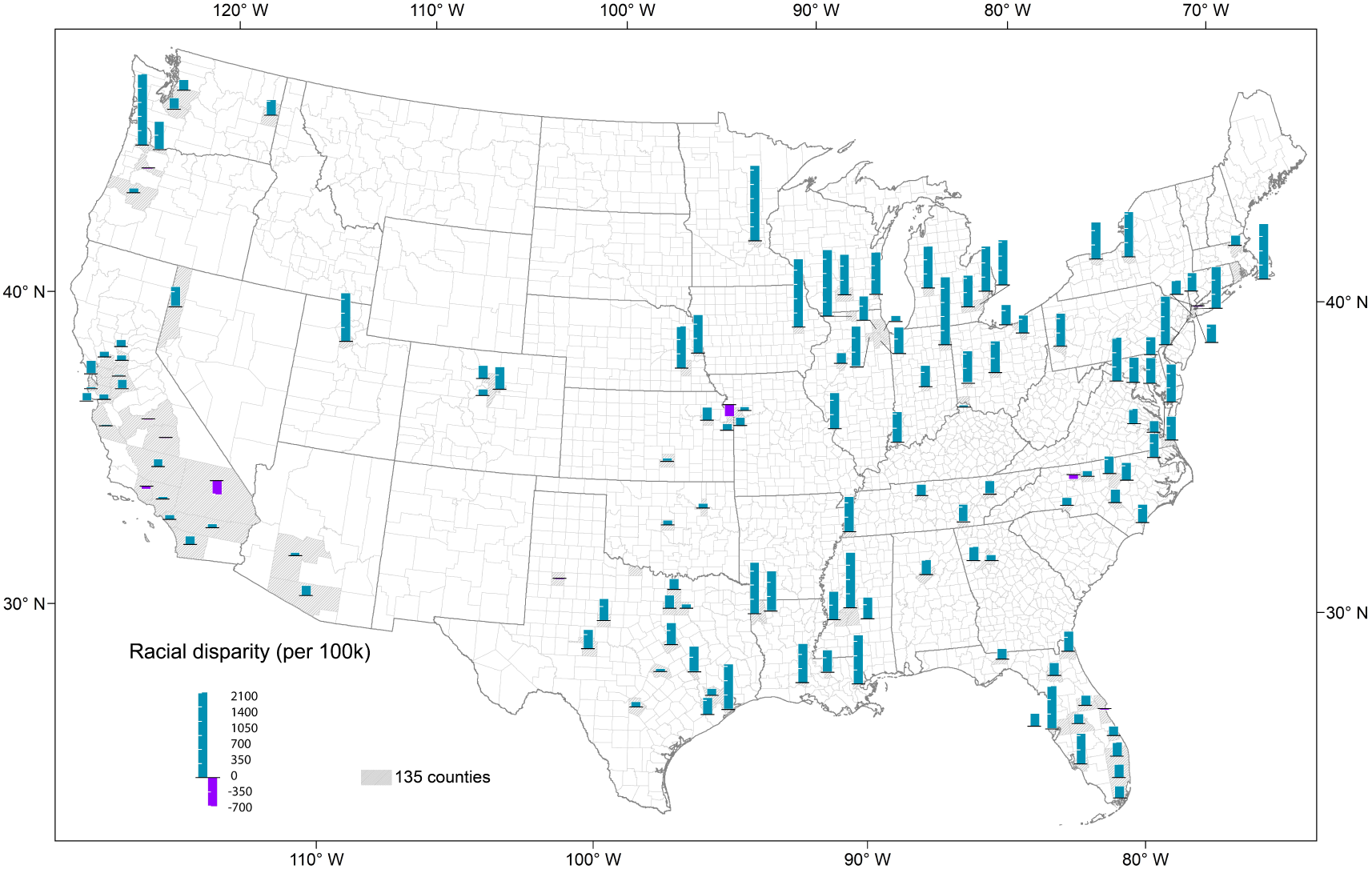
Racial disparity in the SARS-CoV-2 infection rates between black and white individuals in the 135 most urbanized counties of the United States. Cyan column indicates black infection rate is higher than white; purple column indicates white infection rate is higher than black. The height of columns indicates the magnitude of disparity.

**Figure 3.**
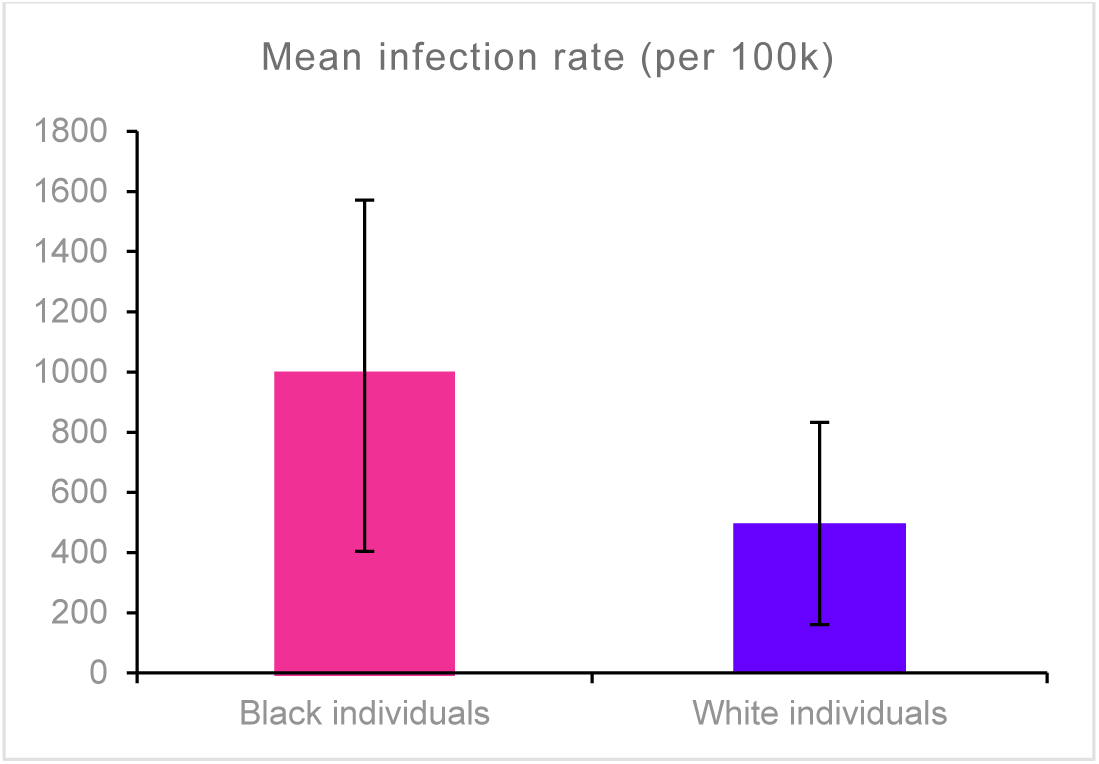
The mean SARS-CoV-2 infection rate of black individuals is approximately twice that of white individuals (988 vs. 497 per 100,000 population). Significant at *p* < 0.001 using a paired *t*-test. The error bar represents standard deviation.

### Characteristics of the sampled counties

Table 1 shows that there are proportionally more white than black people in the study areas, which is consistent with the overall racial composition in the United States. The sample includes an average of 2.6 persons per household and a notably large range in median household income across counties. In terms of the green space factors, the sample includes a high quantity of developed open space and forest cover (Figure 4), and low levels of shrub and scrub, and grassland and herbaceous cover. Figure 4 shows the ratio of four types of green space in each county.

**Figure 4.**
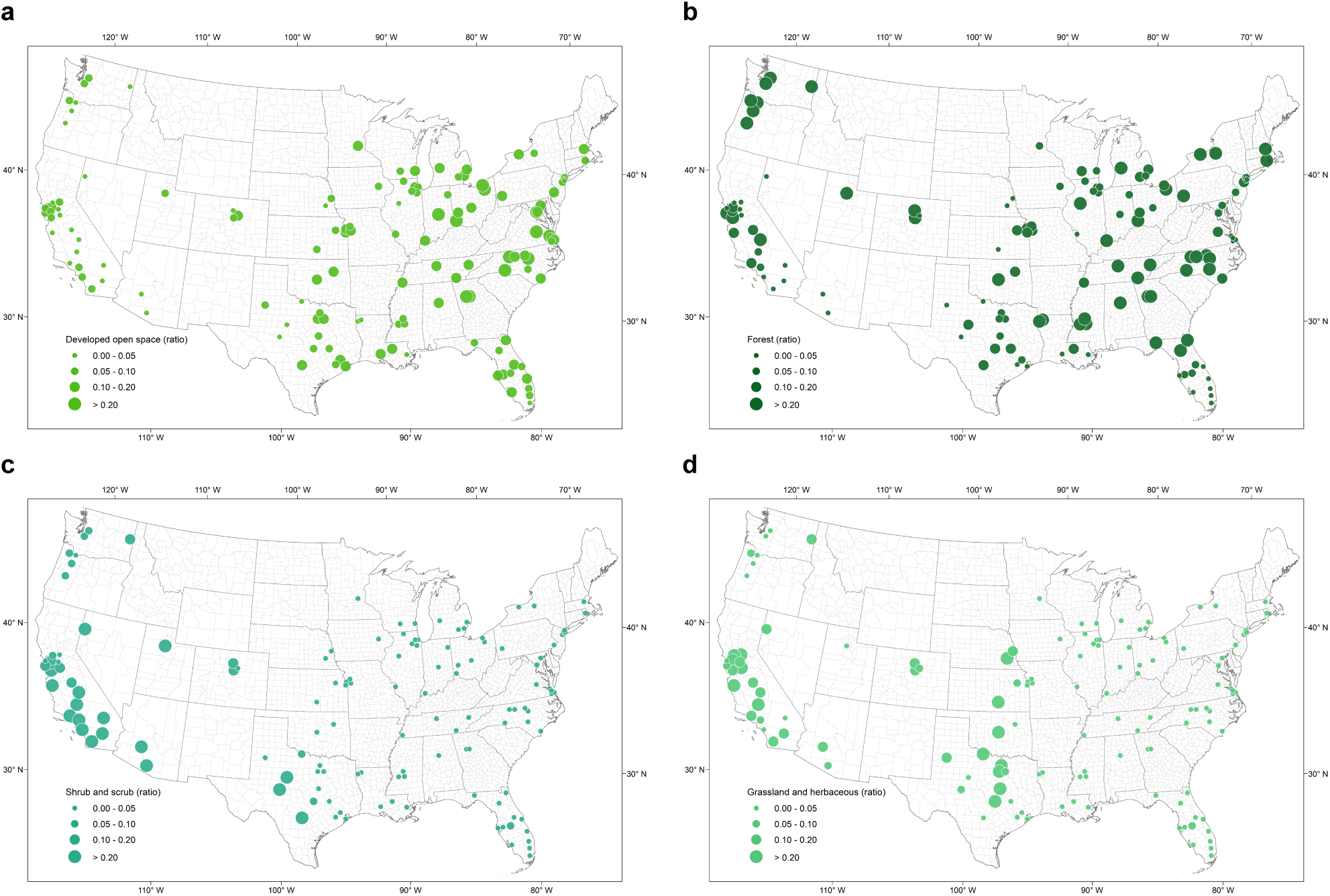
Ratio of four types of green space to total county area, by county. **a** developed open space. **b** forest. **c** shrub and scrub. **d** grassland and herbaceous.

### Modeling association of green spaces and infection disparity

By fitting three hierarchical regression models, we identify three relevant associations (Table 2). Model 1 shows that socioeconomic and demographic factors have a significant association with racial disparity in the SARS-CoV-2 infection rate across the USA’s 135 most urbanized counties (adjusted *R*^2^ of 0.12, *p* = 0.003). We also note that there is a significantly negative association between household size and racial disparity in SARS-CoV-2 infection rates, such that as average household size increases, infection disparity falls.

**Table 2.**
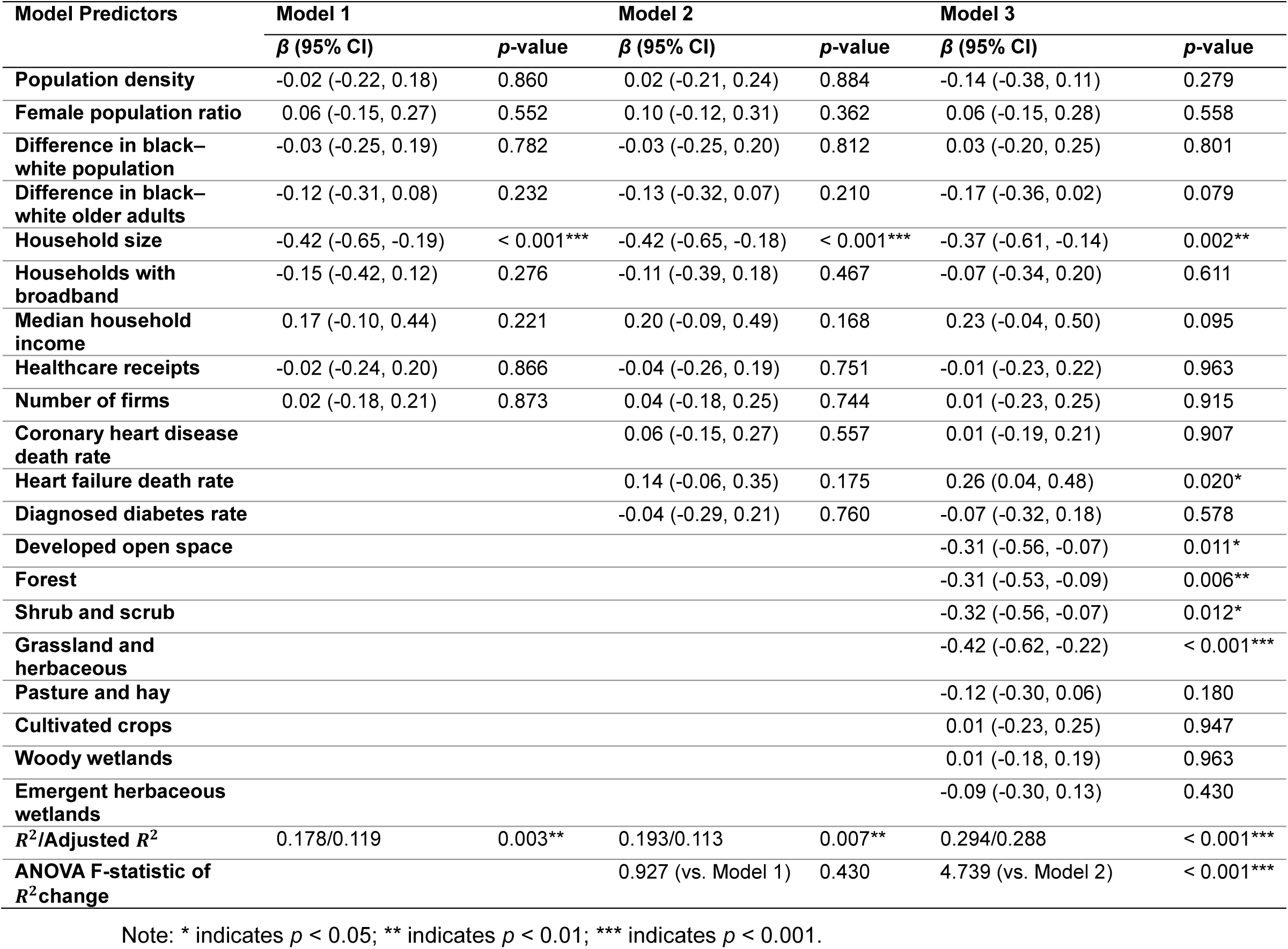
Three hierarchical linear regression results (*N* = 135).

After adding pre-existing chronic disease factors into Model 2, the explanatory power of Model 2 slightly decreases (adjusted *R*^2^ = 0.11, *p* = 0.007). Model 2 shows that the combination of socioeconomic, demographic, and pre-existing chronic disease factors is associated with racial disparity. However, the overall statistical significance of Model 2 was not substantially better than that of Model l (*p* = 0.430 for the sum-of-squares difference).

After adding green space factors into Model 3, the overall explanatory power of Model 3 increased by 18% (adjusted *R*^2^ = 0.29, *p* < 0.001). Model 3 reveals that four green space factors are independently negatively associated with racial disparity. These are: land-cover ratio of developed open space; forest; shrub and scrub; and grassland and herbaceous. In addition, household size is negatively associated with SARS-CoV-2 infection rate disparity between races, while death from heart failure rate is positively associated with it.

To improve our understanding of the associations between the four green spaces and racial disparity in SARS-CoV-2 infection rates, we plotted the independent effects of the four green spaces (using the effects package of the statistical software R^66^). Figure 5 clearly demonstrates the negative relationships between the proportion of the four types of green space in counties and the racial disparity in SARS-CoV-2 infection rates.

**Figure 5.**
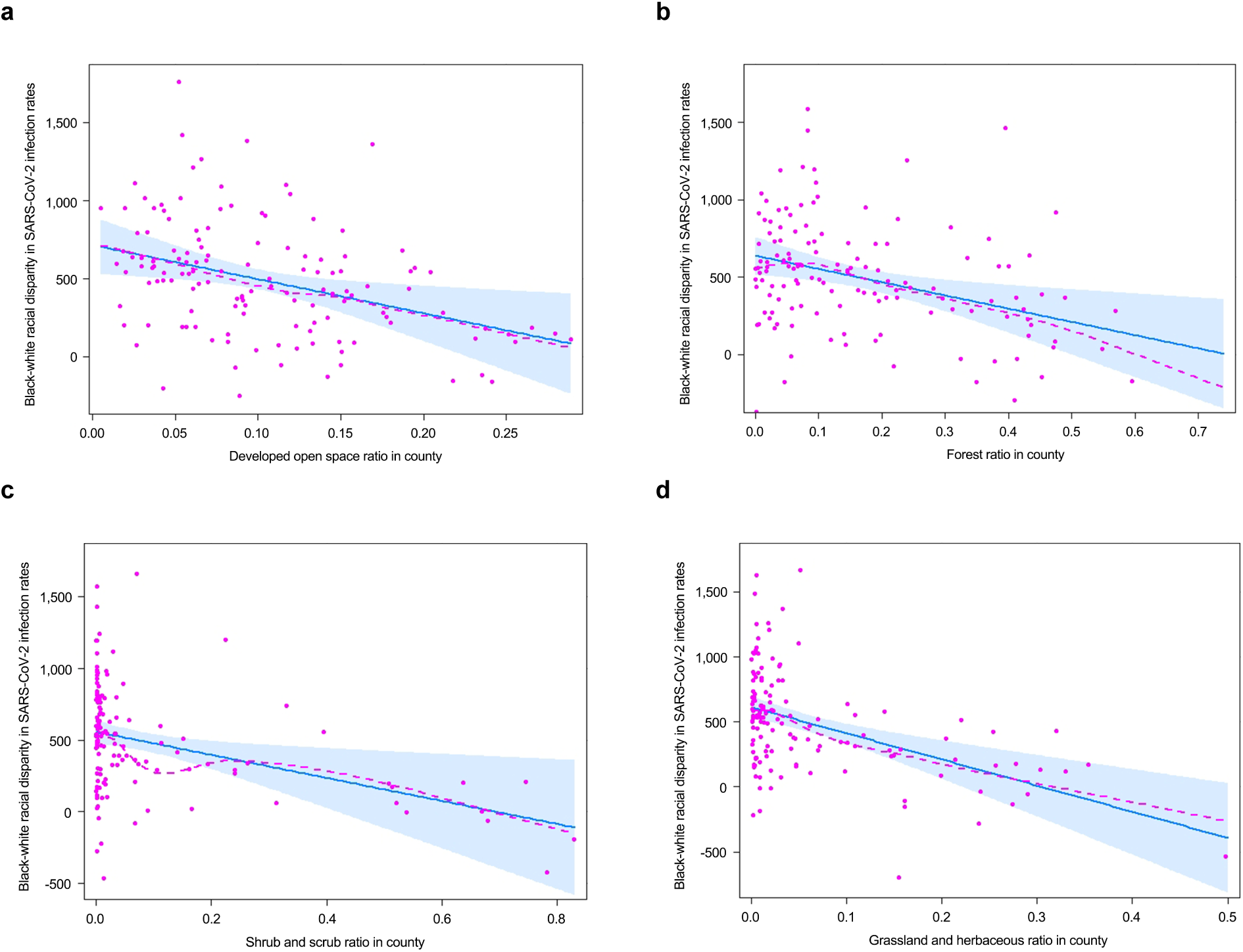
Individual effects of the ratio of four green spaces on the black–white racial disparity in SARS-CoV-2 infection rates (in Model 3, with non-greenspace predictors fixed). **a** developed open space. **b** forest. **c** shrub and scrub. **d** grassland and herbaceous. Shaded areas represent the pointwise 95% confidence interval. Points represent partial residuals. Straight line represents the linear fitting of the effects. Dashed line represents the progressive fitting of the effects.

## Discussion

Significant associations between socioeconomic, demographic, and pre-existing chronic disease factors and the racial disparity in SARS-CoV-2 infection rates have been reported by a number of ecological studies^3, 4, 5, 13, 14^. Some urban environmental factors, such as overcrowding housing conditions, living in senior living communities, living in high-density urban areas, and a long commute distance, are associated with a greater racial disparity in SARS-CoV-2 infection rates ^67, 68, 69, 70, 71^. However, no study examined the relationship between green spaces and racial disparity in SARS-CoV-2 infection rates.

In this study of the 135 urbanized counties in the United States, we found a large and statistically significant disparity in infection rates of the SARS-CoV-2 virus for black and white populations. Using hierarchical linear modelling, we found that the combination of socioeconomic, demographic, and pre-existing chronic disease conditions explained about 11% of variance in racial disparity. Adding green spaces explained an additional 18% of the variance. Four types of green spaces were significantly, negatively associated with the racial disparity in SARS-CoV-2 infection: open space in developed areas, forest, shrub and scrub, and grassland and herbaceous.

In the paragraphs that follow, we consider possible mechanisms through which greater amounts of green spaces within a county might contribute to a reduction in racial disparity in SARS-CoV-2 infection rates. The plausible mechanisms are supported by theoretical and empirical evidence. We elaborate the contributions of our findings and identify questions for future research. We believe this is the first study to find that green space factors have significant independent effects on the racial disparity in COVID-19 infection rates, albeit using an ecological study design.

### Interpretation of key findings

There is growing evidence that the dominant pathway for transmission of the SARS-CoV-2 virus is through aerosol particles ^72, 73, 74^ That is, the primary route of transmission is via virus-containing droplets and aerosols exhaled from infected individuals as they breathe, speak, sing, cough, or sneeze. In indoor settings that lack adequate ventilation, the virus can concentrate in the air, which facilitates its spread. Outdoors, however, because of air movement, and the ease of social distancing, there is a far reduced likelihood of contracting the virus ^75^.

In this study, we found, after controlling for socioeconomic, demographic, pre-exisiting chronic disease, and built-up area factors, that greater proportions of forest, shrub and scrub, grassland, and herbaceous landscapes were each significantly, negatively correlated with the size of discrepancy in black-white infection rates. In urban counties with more green spaces available, the racial disparity in SARS-CoV-2 infection rates was lower than in counties that had less available green space.

How might we account for the findings reported here? We propose four mechanisms that may account for the observed relationships. First, and perhaps most likely, is that green spaces are socio-pedal – that is, they draw people outdoors. A study in Chicago found that individuals using spaces immediately outside apartment buildings were much more likely to be in relatively green spaces than in relatively barren spaces ^76^. A follow-up study of outdoor urban spaces found on average 90% more people used green than barren neighborhood spaces ^77^. Another study reported that urban residents dislike and fear urban neighborhood spaces when they are devoid of vegetation, but that the simple addition of trees and grass was sufficient to transform outdoor common spaces from a space they liked *not at all*, to a space they liked *quite a lot* or *very much* ^78^.

These findings suggest that green neighborhood spaces attract people outdoors. Being outdoors reduces the spread of the virus through two pathways. First, outdoor air movement disperses the virus to levels that are nearly impossible to detect. Second, being outdoors makes it easier than indoors to maintain safe social distance^75^. To the extent that black individuals have disproportionately less access to green spaces that their white counterparts^27, 28, 29, 30^, simply having access to green spaces that pull people outdoors is likely to reduce the racial disparity in infections rates we reported above.

Second, counties with larger portions of green space provided greater access to residents of all racial groups and promoted physical activity before and during the pandemic^21^. This may have enhanced county residents’ immune system regardless of race and moderated factors behind race-based infection disparities^79, 80, 81^. Physical activity conducted in nature provides additional benefits compared with physical activity conducted in indoor environments^82, 83, 84^. Here again, the disproportionate access to green spaces that many black people live with is likely to explain some of the relationships reported above. Black individuals who have access to open green spaces may accrue health benefits from conducting physical activity in these green spaces, and therefore potentially have lower infection rates compared to black people who cannot access open green spaces.

Third, proportionately more green spaces in a county may result in enhanced mental^47^ and social health^48^ regardless of race before and during the pandemic^52^, thus providing multiple benefits that moderate risks that would otherwise fall more heavily on black communities. It is good evidence that, overall, being black in America places heavier stress burdens on individuals than being white, and this distinction may be especially profound during the pandemic^18, 19, 20, 21^. Visual or physical contact with urban green spaces can reduce mental fatigue^50, 85 86^, reduce mental stress^35, 87, 88, 89^, and enhance self-discipline^48^ at the individual level. Such exposure to nature can also reduce negative moods^90, 91^ and verbal and behavioral aggressiveness^49^, which can lead to enhance trust^51, 92^ and collaboration^93^. Taken together, these benefits of exposure to nature can promote immune system function^45^ which may be protective against contracting the virus.

Finally, more green spaces may decrease the SARS-CoV-2 infection risk by improving air quality and decreasing exposure to air pollutants (e.g., PM_2.5_) that are associated with higher SARS-CoV-2 infection rates^94, 95^. Black households tend to be found in higher density, more polluted streets, compared with white households^27, 28, 29, 30^. Thus, the supply of green spaces would be of greater relative benefit to black, rather than white, populations. Studies have shown that air quality in high-density residential areas in urban neighborhoods is significantly worse than that in suburban and rural residential areas^96^. A greater supply of green spaces, especially in urban centers, may reduce this significant environmental disparity^17, 19^, thereby contributing to a lower racial disparity in SARS-CoV-2 infection rates. We note, however, that relying on urban green spaces is not sufficient to create healthy air environments^97^ for the entire urban region. Providing large-scale green spaces beyond the urban core is critical to achieving this goal^98^.

In sum, greater amounts of green spaces may have a stronger health impacts on black individuals and households compared to white individuals and households. This, we suggest, is a crucial mechanism explaining our regression findings. Other things being equal, an increase in the quantity or accessibility of green spaces in an urban area might be expected to have an equal effect on black and white individuals^99, 100^. Black households, however, are less likely to have access green spaces than white households. Thus, the marginal health benefits to black households resulting from more green spaces, will be greater than for whites.

### Contributions and implications

This study makes theoretical and practical contributions, which hope will influence future research, policymaking, and urban design.

First, to the best of our knowledge, this study is the first to measure whether and to what extent green spaces within and beyond developed urban areas are associated with racial disparities in rates of contagious disease infection. Although a few studies have identified some built environment factors, such as crowded living conditions, staying in senior living communities, and dense urban areas, are related to racial disparity in SARS-CoV-2 infection rates^67, 68, 70, 71^, none have addressed green spaces.

Second, we adopted a within-subject research design with a standard spatial sampling unit across the country, giving a representative study for USA urban counties. In contrast with previous studies on COVID-19 that examined associations between SARS-CoV-2 infection rate or COVID-19 mortality rate across counties or cities^10, 101, 102^, our study focused on comparing difference in SARS-CoV-2 infection rates among white and black people in the same county. By comparing racial disparity in infection rates within each county we obtain greater statistical validity, as this approach mitigates bias caused by uneven spread of infections across counties due to differences in national road network accessibility, airports and railway connectivity, governmental regulations, social norms, and quantity and quality of healthcare services.

Third, our study suggests that green space, as a type of urban infrastructure^103^, should be considered a relevant intervention to reduce the racial disparity of infectious diseases with characteristics similar to the current pandemic. Providing an adequate supply of accessible and well-designed green spaces in urban areas, and preserving and developing natural green spaces across counties, is part of an epidemic and pandemic-resilience strategy for highly urbanized areas. Considering that urban and agricultural areas are rapidly encroaching on forest, grassland, and many other natural landscapes inside and/or outside cities worldwide^104, 105^, it is crucial to maintain and increase efforts to preserve scarce urban green spaces. As well as achieving environmental goals, we show that this approach can also be expected to help deliver health and racial equality agenda.

### Limitations and opportunities for future research

This study has some limitations that point to opportunities for future research.

We collected SARS-CoV-2 infection data from county government websites of each of the 135 counties we examined. The quality of data is credible for each county, but the methods of collecting and sorting data clearly varied across counties. Moreover, although there are 229 counties containing or overlapping all cities with a population ≥ 100,000 in the United States, infection data were unavailable for 94 of these counties, leaving a set of high-quality data from 135 counties.

This study only examined racial disparities in SARS-CoV-2 infection rates in the US at the county level. Although the racial disparity in infection rates in the United States might be a good example for countries with diverse racial populations^6, 9, 106^, we suggest that this research should be replicated in other countries, to fully determine the complex issue of racial disparity in infection rates in relation to environmental exposure and pandemic dynamics.

This study provides a base-line analysis of green spaces and racial disparity in pandemic infections. It is limited by the normal constraints of a cross-sectional, ecological design. Cross-sectional associations do not imply causality and we cannot avoid the possibility of ecological fallacy. These are possibilities that can be investigated in further studies. Nevertheless, results from this study echo findings of many published papers that reported causal links between more green spaces, better health status and lower health disparities.

Finally, a caveat can be raised with respect to possible ecological bias. The access to data in SARS-CoV-2 infection rates at the neighborhood scale was not available, which prevented us from examining the distribution of green spaces for races at the neighborhood scale. It is possible an unequal allocation of green spaces could lead to races’ unequal exposures to green spaces, which needs to be further examined in a future study at a finer spatial scale. Two arguments remain reasonable albeit the possibility of that bias. First, more green space leads to greater marginal health benefits for black individuals than white individuals and thus a smaller racial disparity in SARS-CoV-2 infection rates. Second, large patches of natural green spaces outside the city, such as forest and natural grassland and herbaceous lands, can improve the environmental quality of an entire urban area thereby reducing the racial disparity in SARS-CoV-2 infection rates. Future studies should further examine these possibilities.

## Conclusion

This study is an initial effort to understand the relationships between environmental factors and racial disparity in SARS-CoV-2 infection rates. We employed hierarchical regression analysis to perform within-county comparisons of infection rates for black and white individuals. After controlling for socioeconomic, demographic, pre-existing chronic disease, and built-up area factors, we found that greater proportions of forest, shrub and scrub, grassland, and herbaceous landscapes were each significantly, negatively correlated with the size of discrepancy in black-white infection rates. In urban counties with more green spaces, the racial disparity in SARS-CoV-2 infection rates was lower than in counties that had less green space. The findings suggest that the supply of open green space in urban areas and natural green spaces across a county may help to reduce racial disparity in SARS-CoV-2 infection rates. The findings in this study suggest at the potential for green spaces to attenuate racial disparity^55^ and to promote healthier communities.

## Data Availability

1. The infection data were collected from the county health department portals on July 10, 2020.
2. We calculated the SARS-CoV-2 infection rates and the racial disparity for whites and blacks (numbers of cases per 100k) based on the total white and black populations in each county, respectively, which were retrieved from the 2019 census data.
3.We used three datasets to assess green spaces, namely the National Land Cover Datasets in 2016 (NLCD 2016), the Tree Canopy Cover Datasets of the United States Forest Service (USFS), and NDVI.
4. We obtained the demographic and socioeconomic characteristics from the 2019 census data.
5. County-level pre-existing chronic disease factors were collected from the 2016 2018 interactive heart disease and stroke data collated by the Centers for Disease Control and Prevention.

https://www.census.gov/data/tables/time-series/demo/popest/2010s-counties-detail.html

https://www.mrlc.gov/national-land-cover-database-nlcd-2016

https://www.census.gov/programs-surveys/acs/news/updates/2019.html

https://www.cdc.gov/coronavirus/2019-ncov/need-extra-precautions/people-with-medical-conditions.html

https://www.census.gov/quickfacts/orangecountycalifornia

## Author Contributions

BJ proposed the research concept. BJ and YL developed the concept into a full research plan. LC, XML, YWY, WYX conducted the data collection and data analysis under YL’s and BJ’s supervision. YL, BJ and YWY conducted writing of the introduction, discussion, and conclusion. YL, LC, XML, WYX, YWYconducted writing of the methods and results. CW and WCS provided critical revisions for the introduction and discussion.

## Appendix

**Table 1.** The definition for socioeconomic and demographic factors, and pre-existing chronic disease factors.

| Variable Categories | Variables | Item description (formula) | Official definition | VIF Test |
| --- | --- | --- | --- | --- |
| Socioeconomic and demographic factors | Population density | Total population / county area | Population: 2019 county-level census population | Retained |
|  | Female population ratio | Female persons, percent | 2019 county-level census female percent | Retained |
|  | Difference in black-white population | Black population / total population - white population / total population | Black: 2019 county-level census black population<br>White: 2019 county-level census white population | Retained |
|  | Difference in black-white older adults | Older adults of black / black population - older adults of white / white population | Older adults: Age above 65 of the 2019 age group population statistics | Retained |
|  | Household size | Persons per household, 2014-2018 | Persons per household, or average household size, is obtained by dividing the number of persons in households by the number of households (or householders). | Retained |
|  | Housing value | Median value of owner-occupied housing units, 2014-2018 | Specified owner-occupied housing units--one-family houses on less than 10 acres without a business or medical office on the property. | Removed |
|  | Rate of high school graduate or higher | High school graduate or higher, percent of persons age 25 years+, 2014-2018 | High school graduates include people whose highest degree was a high school diploma or its equivalent. These data include only persons 25 years old and over. | Removed |
|  | Rate of households with broadband | Households with a broadband Internet subscription, percent, 2014-2018 | Households with a broadband Internet subscription: The Internet question asked if any member of the household accesses the Internet. | Retained |
|  | Median household income | Median household income (in 2018 dollars), 2014-2018 | Income of households: This includes the income of the householder and all other individuals 15 years old and over in the household, whether they are related to the householder or not. | Retained |
|  | Poverty rate | Persons in poverty, percent (in 2019 census) | Census Bureau uses a set of money income thresholds that vary by family size and composition to determine who is in poverty. If a family's total income is less than the family's threshold, then that family and every individual in it is considered in poverty. | Removed |
| | Healthcare receipts | Total health care and social assistance receipts/revenue, 2012 (\$1,000) / per 100k | The sector includes both health care and social assistance because it is sometimes difficult to distinguish between the boundaries of these two activities. | Retained |
|  | Travel time to work | Mean travel time to work (minutes), workers age 16 years+, 2014-2018 | Travel time to work refers to the total number of minutes that it usually took the person to get from home to work each day during the reference week. | Removed |
|  | Employment rate | Total employment, 2018 / per 100k | Paid employment consists of full- and part-time employees, including salaried officers and executives of corporations. Included are employees on paid sick leave, holidays, and vacations; not included are proprietors and partners of unincorporated businesses. | Removed |
|  | Number of firms | All firms, 2012 / per 100k | Firms equally male-/female-owned, equally minority-/nonminority-owned, and equally veteran-/nonveteran-owned are counted and tabulated as separate categories. | Retained |
| **Pre-existing chronic disease factors** | Coronary heart disease death rate | Coronary heart disease death rate per 100,000 | All ages, all races/ethnicities, both genders, 2016-2018 (not spatial smoothed data) | Retained |
|  | Heart failure death rate | Heart failure death rate per 100,000 | All ages, all races/ethnicities, both genders, 2016-2018 (not spatial smoothed data) | Retained |
|  | Diagnosed diabetes rate | Diagnosed diabetes percentage | Age-adjusted percentage, 20+, 2016 | Retained |
|  | Obesity rate | Obesity percentage | Age-adjusted percentage, 20+. 2016 | Removed |

Note. The detail definition is from the United States Census and Heart disease and stroke data of Centers for Disease Control and Prevention

**Table 2:** Official definition of land-cover types: green space and built-up area factors

| Categories | Classes | Official definition | VIF Test |
| --- | --- | --- | --- |
| **Built-up area factors** | Developed Low Intensity | Areas with a mixture of constructed materials and vegetation. Impervious surfaces account for 20% to 49% percent of total cover. These areas most commonly include single-family housing units. | Removed |
|  | Developed Medium Intensity | Areas with a mixture of constructed materials and vegetation. Impervious surfaces account for 50% to 79% of the total cover. These areas most commonly include single-family housing units. | Removed |
|  | Developed High Intensity | Highly developed areas where people reside or work in high numbers. Examples include apartment complexes, row houses and commercial/industrial. Impervious surfaces account for 80% to 100% of the total cover. | Removed |
| **Green space factors** | Developed open space | Areas with a mixture of some constructed materials, but mostly vegetation in the form of lawn grasses. Impervious surfaces account for less than 20% of total cover. These areas most commonly include large-lot single-family housing units, parks, golf courses, and vegetation planted in developed settings for recreation, erosion control, or aesthetic purposes. | Retained |
|  | Deciduous forest | Areas dominated by trees generally greater than 5 meters tall, and greater than 20% of total vegetation cover. More than 75% of the tree species shed foliage simultaneously in response to seasonal change. | N/A |
|  | Evergreen forest | Areas dominated by trees generally greater than 5 meters tall, and greater than 20% of total vegetation cover. More than 75% of the tree species maintain their leaves all year. Canopy is never without green foliage. | N/A |
|  | Mixed forest | Areas dominated by trees generally greater than 5 meters tall, and greater than 20% of total vegetation cover. Neither deciduous nor evergreen species are greater than 75% of total tree cover. | N/A |
|  | Forest | Combination of deciduous, evergreen, and mixed forest. | Retained |
|  | Shrub and scrub | Areas dominated by shrubs; less than 5 meters tall with shrub canopy typically greater than 20% of total vegetation. This class includes true shrubs, young trees in an early successional stage or trees stunted from environmental conditions. | Retained |
|  | Grassland and herbaceous | Areas dominated by graminoid or herbaceous vegetation, generally greater than 80% of total vegetation. These areas are not subject to intensive management such as tilling, but can be utilized for grazing. | Retained |
|  | Pasture and hay | Areas of grasses, legumes, or grass-legume mixtures planted for livestock grazing or the production of seed or hay crops, typically on a perennial cycle. Pasture/hay vegetation accounts for greater than 20% of total vegetation. | Retained |
|  | Cultivated crops | Areas used for the production of annual crops, such as corn, soybeans, vegetables, tobacco, and cotton, and also perennial woody crops such as orchards and vineyards. Crop vegetation accounts for greater than 20% of total vegetation. This class also includes all land being actively tilled. | Retained |
|  | Woody wetlands | Areas where forest or shrubland vegetation accounts for greater than 20% of vegetative cover and the soil or substrate is periodically saturated with or covered with water. | Retained |
|  | Emergent herbaceous wetlands | Areas where perennial herbaceous vegetation accounts for greater than 80% of vegetative cover and the soil or substrate is periodically saturated with or covered with water. | Retained |

Note: The definition of land classes is derived from The Multi-Resolution Land Characteristics (MRLC) consortium

